# Taxonomic identification and characterization of *Leptospira* from retrospective clinical samples in the Philippines

**DOI:** 10.64898/2026.09.16.26363189

**Authors:** Adeliza Mae L. Realingo, Joanna Ina G. Manalo, Lei Lanna M. Dancel, Kristine Alvarado-Dela Cruz, Arjay Niño A. Digman, Celine Bernice A. Roxas, Desiree D. Argana, Rubelia A. Baterna, Emarld Julian G. Medina, Amalea Dulcene Nicolasora, Timothy John R. Dizon, Francisco Gerardo M. Polotan, Allen Anthony P. Laraño

**Affiliations:** Advanced Molecular Technologies Laboratory, Research Institute for Tropical Medicine, Department of Health, Muntinlupa City, Philippines; Microbiology Department, Research Institute for Tropical Medicine, Department of Health, Muntinlupa City, Philippines

**Author notes:** Correspondence: Adeliza Mae L. Realingo,; Allen Anthony P. Laraño. These authors share senior authorship (Francisco Gerardo M. Polotan, Allen Anthony P. Laraño).

**Keywords:** Leptospirosis, *Leptospira*, mNGS, MAT, qPCR, KrakenUniq

## Abstract

Leptospirosis is a highly endemic neglected tropical disease in the Philippines, where conventional diagnostic methods such as the microscopic agglutination test (MAT) provide limited information on circulating *Leptospira* and are affected by serological cross-reactivity. This study evaluated the utility of shotgun metagenomic next-generation sequencing (mNGS) for pathogen detection and species-level characterization using 20 archived serum-derived total nucleic acid extracts collected from suspected leptospirosis cases. Samples were previously tested by quantitative real-time PCR (qPCR) and MAT, and sequenced using the Illumina MiSeq platform. Host-derived reads were removed using BWA and SAMtools prior to taxonomic profiling with KrakenUniq. Species-level *Leptospira* assignments were retained based on predefined KrakenUniq filtering criteria, including a minimum taxReads threshold and evidence of taxon-specific unique k-mers. *Leptospira* taxReads were detected in 58.8% (n=10/17) of qPCR-positive samples. Notably, mNGS detected pathogenic *Leptospira* in 60% (n=6/10) qPCR-positive but MAT-negative samples, demonstrating its potential to complement conventional serology for etiological characterization. MAT reactivity was predominantly against *L. biflexa* serovar Patoc (n=4/5), with one sample reacting to *L. interrogans* serovar Autumnalis in the MAT-positive samples. Meanwhile, mNGS identified a predominance of *L. interrogans,* with additional detection of *L. borgpetersenii* and *L. kirschneri.* The absence of detectable *Leptospira* in some qPCR-positive samples underscores the sensitivity limitations of mNGS in low-burden or archived samples. Overall, mNGS provided species-level information beyond conventional diagnostics and supports its potential application in *Leptospira* genomic surveillance in the Philippines.

**IMPACT STATEMENT:** Leptospirosis remains an important public health concern in the Philippines, but genomic information on human *Leptospira* infections is limited. Shotgun metagenomic sequencing provided species-level information beyond routine qPCR and MAT. *Leptospira interrogans* predominated, alongside other pathogenic species. These findings establish an initial genomic baseline for *Leptospira* in Philippine clinical specimens and support future surveillance of strain and serovar diversity and transmission.

## INTRODUCTION

Leptospirosis is a zoonotic disease caused by pathogenic *Leptospira* species, with clinical manifestations ranging from mild flu-like symptoms, including myalgia, conjunctivitis, and gastrointestinal symptoms, to severe disease involving multiorgan damage (Amilasan et al., 2012; Ji et al., 2023; Lu et al., 2022). Severe cases may result in jaundice, renal failure, pulmonary hemorrhage, acute respiratory distress syndrome, and other life-threatening conditions (Amilasan et al., 2012; Chen et al., 2021; Ji et al., 2023). Early and accurate diagnosis is critical for preventing disease progression and improving patient outcomes; however, the nonspecific clinical presentation of leptospirosis makes diagnosis challenging, particularly in resource-limited settings (Caimi et al., 2012; Mendoza & Rivera, 2021).

In the Philippines, *Leptospira* has been detected in both animal reservoirs and environmental sources, particularly in areas affected by flooding and heavy rainfall. Villanueva et al. (2010) reported high seroprevalence among rats collected in Metro Manila and Laguna and identified *L. interrogans* serovars Manilae and Losbanos, serogroup Grippotyphosa, and *L. borgpetersenii* serogroup Javanica among the isolates. These findings demonstrate the circulation of diverse pathogenic *Leptospira* in Philippine reservoirs. However, molecular characterization of *Leptospira* from human clinical specimens remains limited, as much of the available molecular evidence has focused on animal and environmental sources (Yanagihara et al., 2007; Mendoza & Rivera, 2019). This limits our understanding of which pathogenic *Leptospira* species and lineages are circulating among human infections and constrains the development of genomic surveillance strategies in the country.

The *Leptospira* genus comprises substantial species and serovar diversity, including pathogenic, intermediate, and saprophytic lineages (Vincent et al., 2019; Korba et al., 2021). More recent genomic classifications have further divided these lineages into P1 and P2 pathogenic groups and S1 and S2 saprophytic groups, reflecting increasing recognition of genomic diversity within the genus (Vincent et al., 2019). The distinction among pathogenic species and lineages is epidemiologically important because different *Leptospira* lineages may vary in their geographic distribution, host associations, and clinical relevance. Consequently, identifying the species circulating in human infections represents an important step toward understanding the molecular epidemiology of leptospirosis (Jayasundara et al., 2021; Boonsilp et al., 2013; Mendoza & Rivera, 2021).

Culture remains a conventional method for identifying *Leptospira* but it is time-consuming and has limited utility for timely clinical diagnosis. Serological methods, particularly the microscopic agglutination test (MAT), remain widely used and are considered an important reference method for leptospirosis diagnosis (Goris & Hartskeerl, 2014). However, MAT is of limited value during early infection because antibodies typically become detectable only several days after exposure (Chen et al., 2021; Ahmed et al., 2009). Its interpretation may also be complicated by cross-reactivity among serovars, variation in laboratory procedures, and host immune responses, limiting accurate identification of the infecting serovar (Markovich et al., 2012; Lu et al., 2022). Indeed, MAT correctly predicted the infecting serovar in only 33% of human cases in one study in Thailand when compared with cross-agglutinin absorption testing (Smythe et al., 2009). Its sensitivity also remains limited even when combined with culture (Limmathurotsakul et al., 2012). Thus, a negative serological result, particularly during acute disease, does not exclude infection (Sykes et al., 2022b).

Nucleic acid amplification tests provide more specific detection of *Leptospira* during the acute phase of infection, when bacterial DNA may be detectable in blood, particularly before or early in antibiotic treatment (Ahmed et al., 2009; Thaipadunpanit et al., 2011; Manalo et al., 2026). Ahmed et al. (2009) developed and validated a SYBR Green-based real-time PCR targeting the *secY* gene, which has subsequently been adapted by the Microbiology Department of the Research Institute for Tropical Medicine (RITM) for the detection of pathogenic *Leptospira* (Manalo et al., 2026).

Despite the substantial burden of leptospirosis in the Philippines, genomic information on *Leptospira* circulating among human infections remains limited. Current diagnostic approaches provide complementary but incomplete information: the qPCR assay based on Ahmed et al. (2009) and currently used provides targeted detection of pathogenic *Leptospira* but does not resolve the infecting species, serovar, or genetic lineage, while MAT provides serological information but may be affected by cross-reactivity and paradoxical reactions. Consequently, there is a need for approaches that can detect *Leptospira* while also providing information on the diversity of pathogenic species circulating among human infections in the country.

Emerging genomic approaches, including shotgun metagenomic next-generation sequencing (mNGS), offer an opportunity to address these limitations by providing culture-independent taxonomic information directly from clinical specimens (Sharpton et al., 2014). Unlike targeted PCR assays, shotgun mNGS does provide a broader, agnostic assessment of microbial sequences present in a specimen (Sharpton et al., 2014; Shi et al., 2022; Jiang et al., 2022). With sufficient pathogen genome recovery, genomic sequencing can further support species-level characterization, epidemiological surveillance, and investigation of potential transmission links between human infections and animal or environmental reservoirs (Lehmann et al., 2014; Sykes et al., 2022b; Grillova et al., 2023).

In this study, we evaluated shotgun mNGS on archived serum-derived total nucleic acid extracts for the detection and species-level characterization of *Leptospira.* Shotgun mNGS was selected for its unbiased, agnostic capacity to detect potentially unexpected *Leptospira* taxa without requiring prior knowledge of circulating species or serovars. Taxonomic profiling was performed using KrakenUniq, which incorporates taxon-specific unique k-mers to provide additional evidence for taxonomic assignments and is therefore useful for evaluating low-abundance pathogen signals in host-dominated clinical specimens (Breitwieser et al., 2018). This study provides a baseline for species-level *Leptospira* surveillance in human clinical samples and establishes a foundation for future genomic investigations in the country.

## MATERIALS AND METHODS

### A. Sample acquisition

All laboratory methods and computational resources for data analysis were performed at the Advanced Molecular Technologies Laboratory (AMTL) of the Research Institute for Tropical Medicine (RITM). Archived total nucleic acid (TNAs) extracts derived from serum samples, using QIAamp DNA Blood Mini Kit, of patients with suspected leptospirosis were obtained from the Microbiology Department of RITM and included in this study through convenience sampling. The archived extracts originated from samples collected between 2018 and 2024 and had previously undergone diagnostic testing using microscopic agglutination test (MAT) and quantitative polymerase chain reaction (qPCR).

Of the 57 archived TNA extracts available for consideration, 20 were selected for downstream processing based on the following inclusion criteria: (1) availability of sufficient TNA extract for testing; (2) a calculated total DNA input exceeding 9.7 ng based on Qubit fluorometric quantification and the available extract volume; and (3) availability of corresponding diagnostic qPCR and MAT results. Nucleic acid extracts that did not meet the minimum DNA input requirement were excluded because the available nucleic acid was insufficient for downstream analyses. Thus, the final sample size of 20 was determined by the availability and quality of the archived extracts rather than by prospective sample size calculation.

Among the 20 selected extracts, five were positive by both qPCR and MAT, 10 were positive by qPCR but negative by MAT, two were positive by qPCR with no MAT result available, and three were negative by both qPCR and MAT. These archived diagnostic results were retained to describe the laboratory classification of the selected samples. Unique study identifiers were assigned to all extracts to maintain confidentiality and prevent patient linkage.

### B. DNA purification, library preparation, and sequencing

The DNA samples were prepared using Agencourt AMPure XP, PCR Purification before library preparation. A total of 30 µL was aliquoted from the archived total nucleic acid extracts in a microtube plate. Following the required 0.9X AMPure:Sample volume ratio, 27 µL of AMPure XP was added to each sample. After purification steps, the supernatant was collected and transferred to a labeled new plate. The concentration of the samples was quantified (in ng/µL) using Qubit™ dsDNA High Sensitivity Assay Kit (Thermo Fisher Scientific, USA). The total DNA input was normalized based on the 30 µL input per sample. The purified DNA for each sample was then subjected to library preparation using the protocol Illumina DNA Prep Kit (Reference Guide, Document # 1000000025416 v10). We loaded a total of 600 µL of prepared sequencing libraries, spiked with 10% PhiX control, onto a MiSeq v3 reagent cartridge. A no-template control (NTC) was processed through the same library preparation protocol and included in the sequencing run to monitor for reagent and sequencing contamination. Sequencing was performed on the Illumina MiSeq for 36 hours, after which raw paired-end reads were retrieved for downstream analysis.

### C. Bioinformatics and Data Analysis

#### Host read removal

To remove host-derived sequences, raw sequencing reads were aligned against the human reference genome GRCh38.p14 using BWA v0.7.19-r1273. The resulting alignment files were processed using SAMtools v1.3.1 to identify reads that mapped to the human reference genome. Reads mapping to the human genome were excluded, and the unmapped reads were retained as host-depleted reads for downstream taxonomic analysis.

#### Taxonomic profiling using KrakenUniq

Taxonomic profiling was performed on the host-depleted reads using KrakenUniq v1.0.4 and the KrakenUniq Microbial2025 database (https://benlangmead.github.io/aws-indexes/k2#krakenuniq). The host-depleted reads were classified by the microbial database and generated taxonomic assignment reports. We used the KrakenUniq predefined filtering criteria of ≥10 taxReads and ≥1,000 unique k-mers to ensure high-confidence taxonomic assignments (Breitwieser et al., 2018). Taxonomic profiles were subsequently summarized and visualized using custom R scripts (https://github.com/lanadelrea/PH-Leptospirosis-Analyses).

## RESULTS

### KrakenUniq detection and taxonomic classification of *Leptospira*

A total of 20 archived serum samples were analyzed by shotgun metagenomic sequencing and compared with conventional diagnostic results from qPCR and MAT (Table 1). Of the 20 samples, 85.0% (n=17/20) were qPCR-positive and 15.0% (n=3/20) were qPCR-negative. MAT was performed for 18 samples, of which 27.8% (n=5/18) were MAT-positive and 72.2% (n=13/18) were MAT-negative. Among the five MAT-positive samples, 80.0% (n=4/5) were identified as *Leptospira biflexa* serovar Patoc, while one sample (20.0%, n=1/5; LS24-098) was identified as *L. interrogans* serovar Autumnalis. MAT was not performed for LS24-002 and LS24-099 because of insufficient serum volume.

**Table 1.**
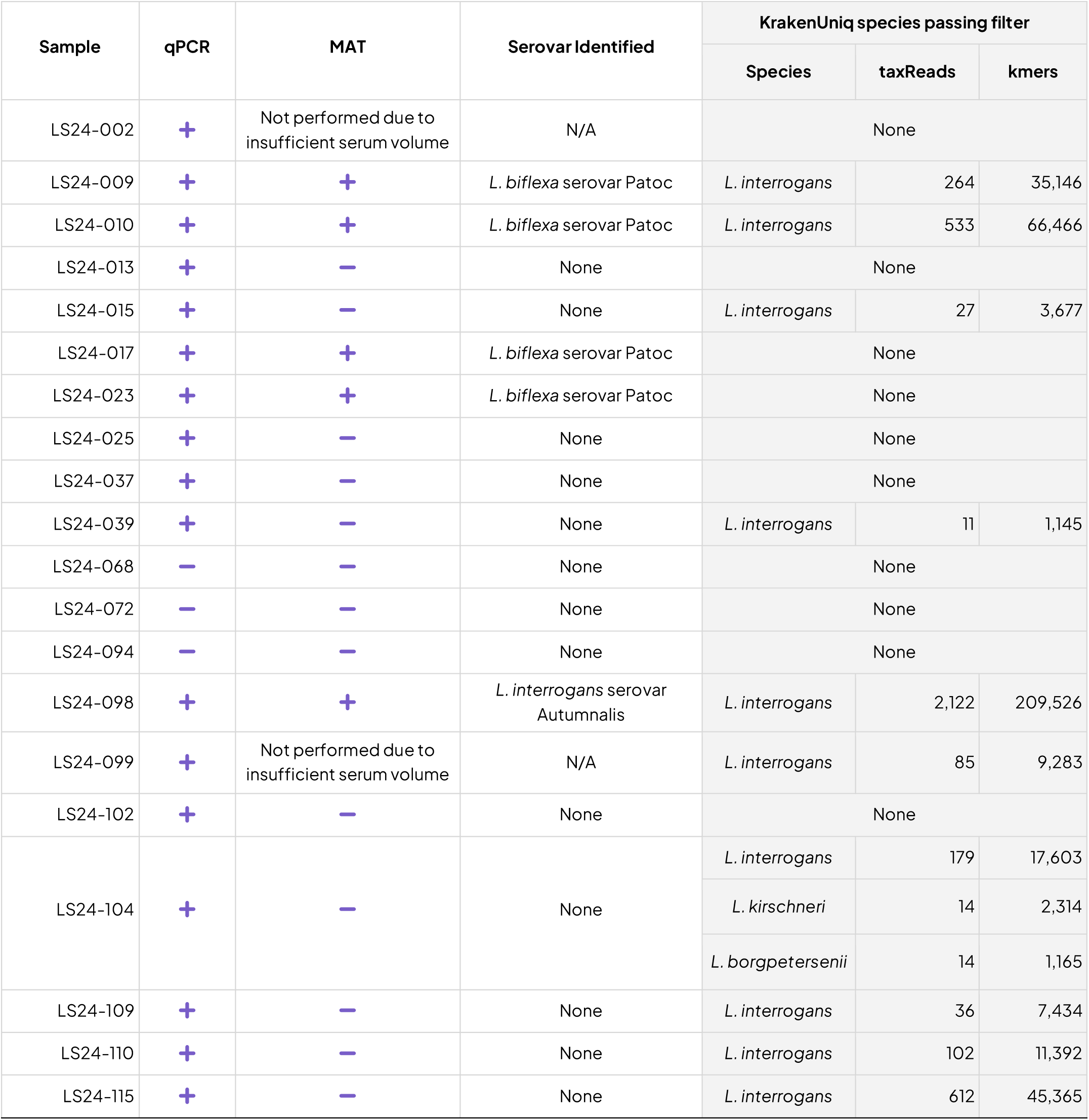
Diagnostic results and high-confidence *Leptospira* assignments by KrakenUniq.

Using the predefined KrakenUniq filtering criteria, high-confidence *Leptospira* assignments were identified in 50.0% (n=10/20) of all samples and in 58.8% (n=10/17) of qPCR-positive samples (Table 1). Among qPCR-positive/MAT-positive samples, *Leptospira* was detected in 60.0% (n=3/5; LS24-009, LS24-010, and LS24-098) (Table 2). Among qPCR-positive/MAT-negative samples, high-confidence *Leptospira* assignments were detected in 60.0% (n=6/10) (Table 3). One of the two qPCR-positive samples for which MAT was not performed, LS24-099, yielded a high-confidence *L. interrogans* assignment (Table 4). None of the qPCR-negative/MAT-negative samples yielded a *Leptospira* assignment meeting the filtering criteria (Table 5).

**Table 2.** KrakenUniq detection of *Leptospira* in qPCR-positive and MAT-positive samples.

| Sample | qPCR | MAT | Serovar Identified | KrakenUniq species passing filter |
| --- | --- | --- | --- | --- |
| LS24-009 | + | + | *L. biflexa* serovar Patoc | *L. interrogans* |
| LS24-010 | + | + | *L. biflexa* serovar Patoc | *L. interrogans* |
| LS24-017 | + | + | *L. biflexa* serovar Patoc | None |
| LS24-023 | + | + | *L. biflexa* serovar Patoc | None |
| LS24-098 | + | + | *L. interrogans* serovar Autumnalis | *L. interrogans* |

**Table 3.**
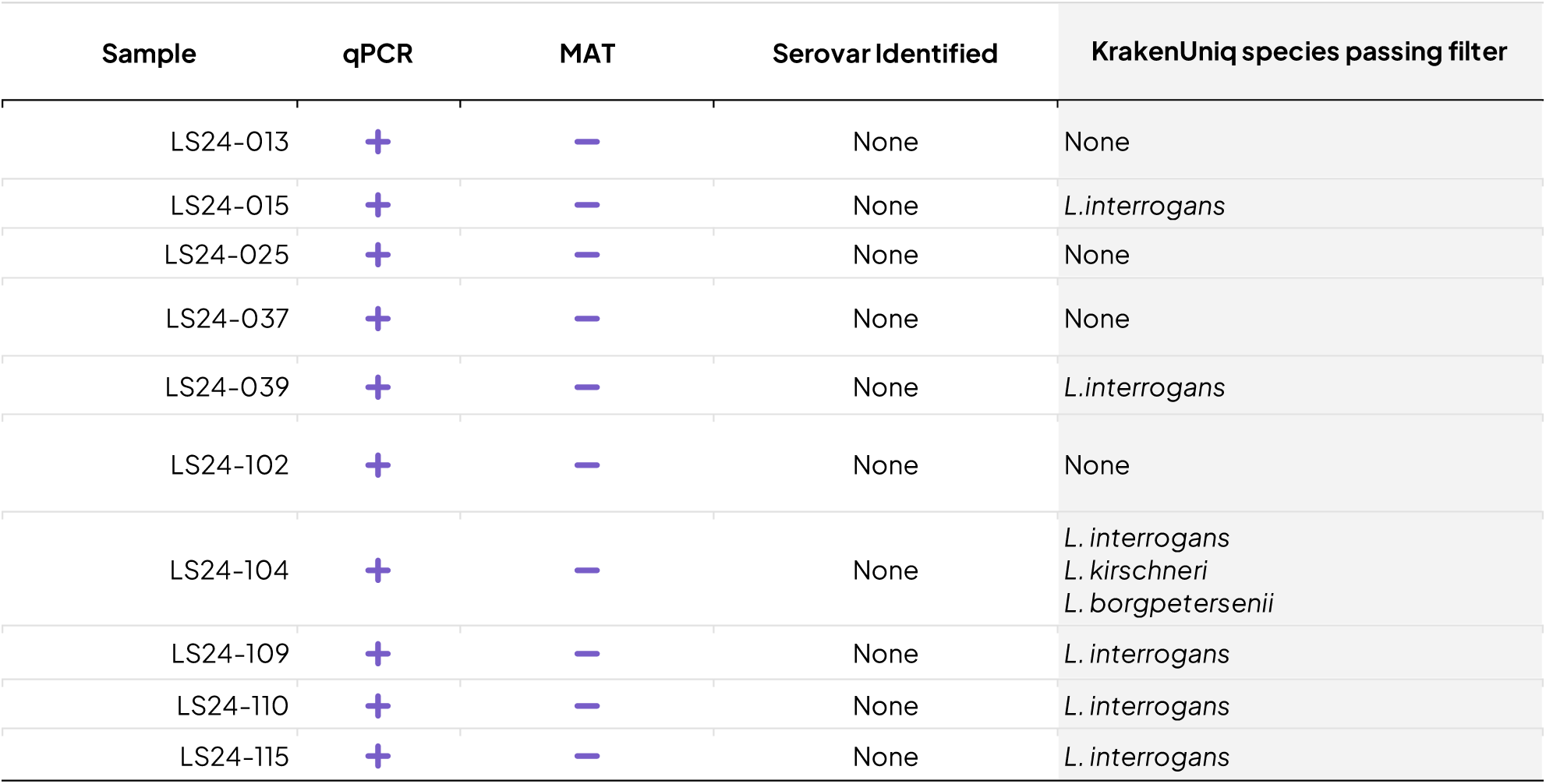
KrakenUniq detection of *Leptospira* in qPCR-positive and MAT-negative samples.

| Sample | qPCR | MAT | Serovar Identified | KrakenUniq species passing filter |
| --- | --- | --- | --- | --- |
| LS24-013 | + | — | None | None |
| LS24-015 | + | — | None | *L. interrogans* |
| LS24-025 | + | — | None | None |
| LS24-037 | + | — | None | None |
| LS24-039 | + | — | None | *L. interrogans* |
| LS24-102 | + | — | None | None |
| LS24-104 | + | — | None | *L. interrogans*<br>*L. kirschneri*<br>*L. borgpetersenii* |
| LS24-109 | + | — | None | *L. interrogans* |
| LS24-110 | + | — | None | *L. interrogans* |
| LS24-115 | + | — | None | *L. interrogans* |

**Table 4.** qPCR-positive and no MAT samples and the taxonomic classification of *Leptospira* reads.

| Sample | qPCR | MAT | Serovar Identified | KrakenUniq species passing filter |
| --- | --- | --- | --- | --- |
| LS24-002 | + | Not performed due to insufficient serum volume | N/A | None |
| LS24-099 | + | Not performed due to insufficient serum volume | N/A | *L. interrogans* |

**Table 5.** qPCR-negative and MAT-negative samples.

| Sample | qPCR | MAT | Serovar Identified | KrakenUniq species passing filter |
| --- | --- | --- | --- | --- |
| LS24-068 | - | - | None | None |
| LS24-072 | - | - | None | None |
| LS24-094 | - | - | None | None |

*L. interrogans* was the predominant species among the high-confidence assignments. The highest taxReads count was observed in LS24-098, with 2,122 taxReads and 209,526 unique k-mers. This sample was positive by both qPCR and MAT and was identified by MAT as *L. interrogans* serovar Autumnalis. Other samples with comparatively high *L. interrogans* taxReads included LS24-115 (612 taxReads), LS24-010 (533 taxReads), LS24-009 (264 taxReads), and LS24104 (179 taxReads). LS24-104 also yielded high-confidence assignments to *L. kirschneri* (14 taxReads; 2,314 unique k-mers) and *L. borgpetersenii* (14 taxReads; 1,165 unique k-mers).

Notably, high-confidence *Leptospira* assignments were recovered from 60.0% (n=6/10) of qPCR-positive/MAT-negative samples, providing species-level taxonomic information in specimens without detectable MAT reactivity. LS24-104 showed the greatest species diversity, with assignments to *L. interrogans, L. kirschneri,* and *L. borgpetersenii.* LS24-099, for which MAT was not performed because of insufficient serum volume, also yielded a high-confidence *L. interrogans* assignment (85 taxReads; 9,283 unique k-mers).

### Other species-level taxonomic assignments

In addition to *Leptospira,* several non-*Leptospira* species-level assignments met the same KrakenUniq filtering criteria (Figure 2). These included *Cutibacterium acnes, Cutibacterium modestum, Acinetobacter ursingii, Acinetobacter haemolyticus, Myroides odoratimimus, Stutzerimonas stutzeri, Enterobacter hormaechei, Enterococcus faecalis,* and *Methylorubrum populi.* The greatest number of non-*Leptospira* assignments was observed in LS24-010, where *A. ursingii* (140 taxReads; 28,265 unique k-mers), *M. odoratimimus* (115 taxReads; 8,839 unique k-mers), and *C. acnes* (10 taxReads; 1,095 unique k-mers) were detected alongside *L. interrogans* (533 taxReads; 66,466 unique k-mers). LS24-013 yielded *C. acnes* (55 taxReads; 4,952 unique k-mers) and *C. modestum* (18 taxReads; 1,351 unique k-mers), while LS24-099 yielded *E. hormaechei* (92 taxReads, 3,245 unique k-mers) and *E. faecalis* (32 taxReads; 1,920 unique k-mers) alongside *L. interrogans* (85 taxReads; 9,283 unique k-mers). Other non-*Leptospira* assignments included *S. stutzeri* in LS24-094, *A. haemolyticus* in LS24-102, and *Methylorubrum populi* in LS24-104 (Supplementary Table 1).

Among samples with high-confidence *Leptospira* assignments, *L. interrogans* generally constituted the predominant species-level assignment. In LS24-009, LS24-015, LS24-039, LS24-098, LS24-109, LS24-110, and LS24-115, *Leptospira* accounted for 95.7–100% of filtered non-host taxReads (Figure 1; Figure 2). In contrast, non-*Leptospira* taxa comprised a larger proportion of filtered assignments in samples such as LS24-010, where *L. interrogans* represented 66.8% of filtered non-host taxReads (Figure 1; Supplementary Table 1).

**Figure 1.**
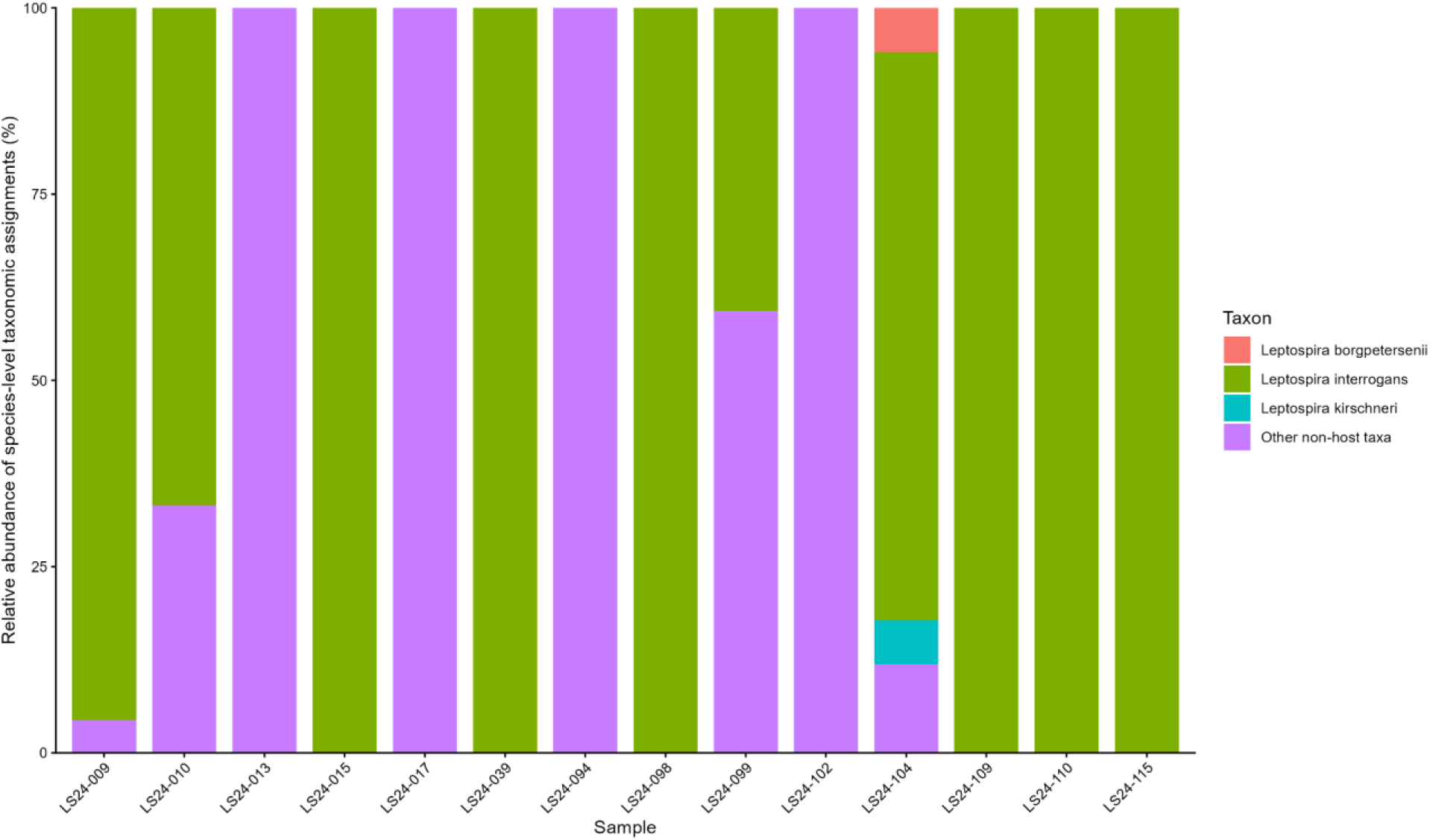
Relative abundance of *Leptospira* and other species-level taxonomic assignments

Moreover, the non-*Leptospira* assignments were observed in both *Leptospira-*positive and *Leptospira-*negative samples. These findings demonstrate that the shotgun metagenomic workflow recovered a range of species-level non-host taxonomic assignments from clinical specimens in addition to *Leptospira.* However, these assignments were not independently evaluated for clinical significance and should therefore be interpreted as metagenomic taxonomic classifications rather than evidence of concomitant infection. Finally, the NTC yielded no reads, providing no evidence of reagent or sequencing contamination in the run.

## DISCUSSION

Metagenomic sequencing of clinical samples for *Leptospira* detection and characterization remains challenging because pathogen DNA is typically present at very low abundance relative to host-derived nucleic acids. Previous studies have reported extremely low proportions of *Leptospira* sequences in clinical specimens, ranging from 0.016% of total reads in cerebrospinal fluid from a patient with neuroleptospirosis (Wilson et al., 2014) to only 0.0008%-0.15% of reads mapped to *Leptospira* reference genomes in clinical specimens analysed by standard metagenomic sequencing without enrichment (Grillova et al. 2023). Similarly, Chen et al. (2021) reported genome coverage of only 3.9 x 10^-3^ % from four reads in a case of severe pulmonary hemorrhagic leptospirosis. This low pathogen burden, combined with the overwhelming proportion of host-derived nucleic acids, can substantially reduce the effective sequencing depth available for *Leptospira* detection and genomic characterization (Grillova et al., 2023).

Consistent with these challenges, we observed substantial variability in *Leptospira* detection across serum samples. Application of the KrakenUniq filtering criteria of ≥10 taxReads and ≥1,000 unique k-mers resulted in high-confidence *Leptospira* assignments in 58.8% (n=10/17) of qPCR-positive samples, including 60.0% (n=6/10) of qPCR-positive/MAT-negative samples. Several qPCR-positive samples therefore yielded either no *Leptospira* reads or signals below the filtering threshold, highlighting the limited sensitivity of untargeted metagenomic sequencing when pathogen DNA is present at low concentrations. These findings indicate that although mNGS can recover *Leptospira* sequences from clinical specimens, the high host-to-pathogen DNA ratio remains a major constraint on pathogen detection and genome recovery (Grillova et al., 2023).

The absence of *Leptospira* reads, or failure to meet the filtering criteria, in some qPCR-positive samples should not necessarily be interpreted as evidence of the absence of pathogen DNA (Grillova et al., 2023). qPCR generally provides greater analytical sensitivity for detecting low concentrations of target nucleic acid than untargeted metagenomic sequencing because amplification enables detection of targets that may be represented by very few or no sequencing reads (Sykes et al., 2022b; Grillova et al., 2023). In addition, the use of archived total nucleic acid extracts stored for several years may have reduced sequencing performance, as prolonged cryopreservation of plasma has been associated with lower detectable yield (Grundy et al., 2023; Yuwono et al., 2022). Thus, the discordance observed between qPCR and metagenomic detection likely reflects differences in analytical sensitivity and sample quality rather than a direct contradiction between the methods (Spatz & Afonso, 2024; Grillova et al., 2023).

The choice of taxonomic classification method and reference database is an important consideration in metagenomic analysis, particularly for low-abundance pathogens (Breitwieser et al., 2018; Wood et al., 2019). Recent benchmarking studies have shown that larger and more diverse reference databases can improve taxonomic resolution and classification performance, although computational efficiency remains an important consideration as reference repositories continue to expand (Breitwieser et al., 2018; Wood et al., 2019; Piro & Reinert, 2023). In this study, KrakenUniq was used to characterize the taxonomic composition of the sequencing reads because its classification approach incorporates taxon-specific unique k-mers as additional evidence for taxonomic assignments (Breitwieser et al., 2018). This feature is particularly relevant to low-abundance pathogen detection, where short or conserved sequence matches may otherwise result in ambiguous assignments in complex, host-dominated samples (Breitwieser et al., 2018; Wood et al., 2019). We further applied the filtering thresholds of ≥10 taxReads and ≥1,000 unique k-mers to identify species-level assignments with stronger taxonomic support. The use of species-level taxReads provided a more conservative measure of taxonomic support than total reads assigned to a taxon and revealed a predominance of *L. interrogans* among samples meeting the filtering criteria. Additional assignments to *L. borgpetersenii* and *L. kirschneri* were observed in LS24-104.

The same filtering approach also identified several non-*Leptospira* species-level assignments meeting the predefined thresholds (Figure 2). These included *Cutibacterium acnes, C. modestum, Acinetobacter ursingii, A. haemolyticus, Myroides odoratimimus, Stutzerimonas stutzeri, Enterobacter hormaechei, Enterococcus faecalis,* and *Methylorubrum populi.* Their detection indicates that the filtering strategy retained taxonomically supported non-*Leptospira* assignments rather than selectively identifying *Leptospira*. Some of these organisms are associated with human skin, environmental sources, or other microbial niches, and their detection in metagenomic datasets may represent background microbial DNA, specimen-associated microorganisms, or laboratory or sequencing contamination (Lusk, 2014; Simner et al., 2018). The presence of non-*Leptospira* taxa therefore emphasizes the importance of interpreting species-level metagenomic classifications in the context of specimen type, abundance, sequencing background, and controls (Zinter et al., 2019; Bihl et al., 2022). Although the use of unique k-mer and taxRead thresholds provide additional support for taxonomic classification, these criteria alone do not establish organism viability or clinical relevance (Simner et al., 2018). A no-template control (NTC) included in the sequencing run yielded no reads, indicating that the non-*Leptospira* signals did not derive from detectable reagents or sequencing contamination (Salter et al., 2014; Eisenhower et al., 2019), and supporting the interpretation that the non-*Leptospira* signals did not originate from the sequencing workflow. Nonetheless, these findings should not be interpreted as evidence of concurrent infection or clinical causation.

**Figure 2.**
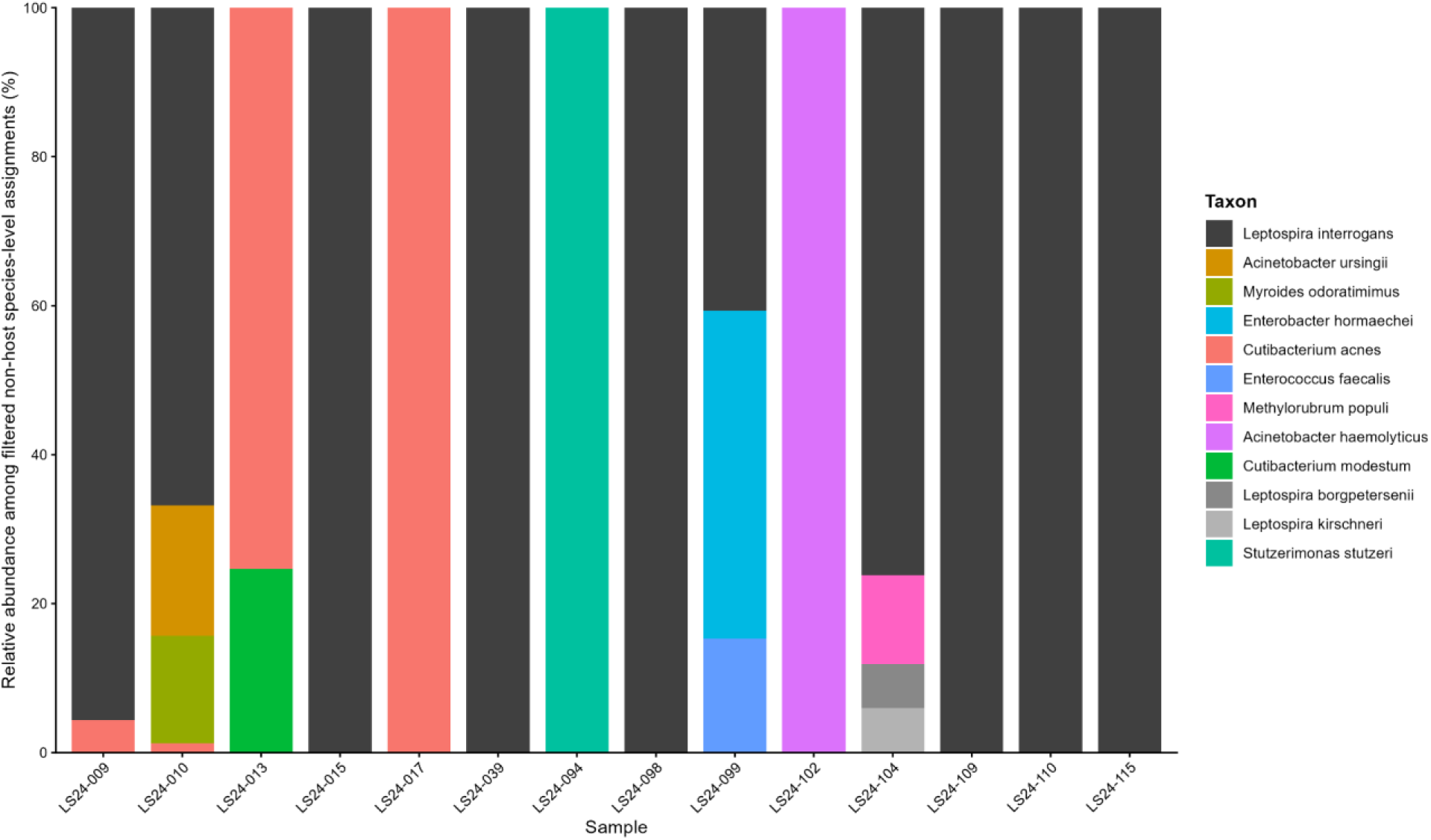
Species-level taxa passing the KrakenUniq filtering criteria

**Figure 3.**
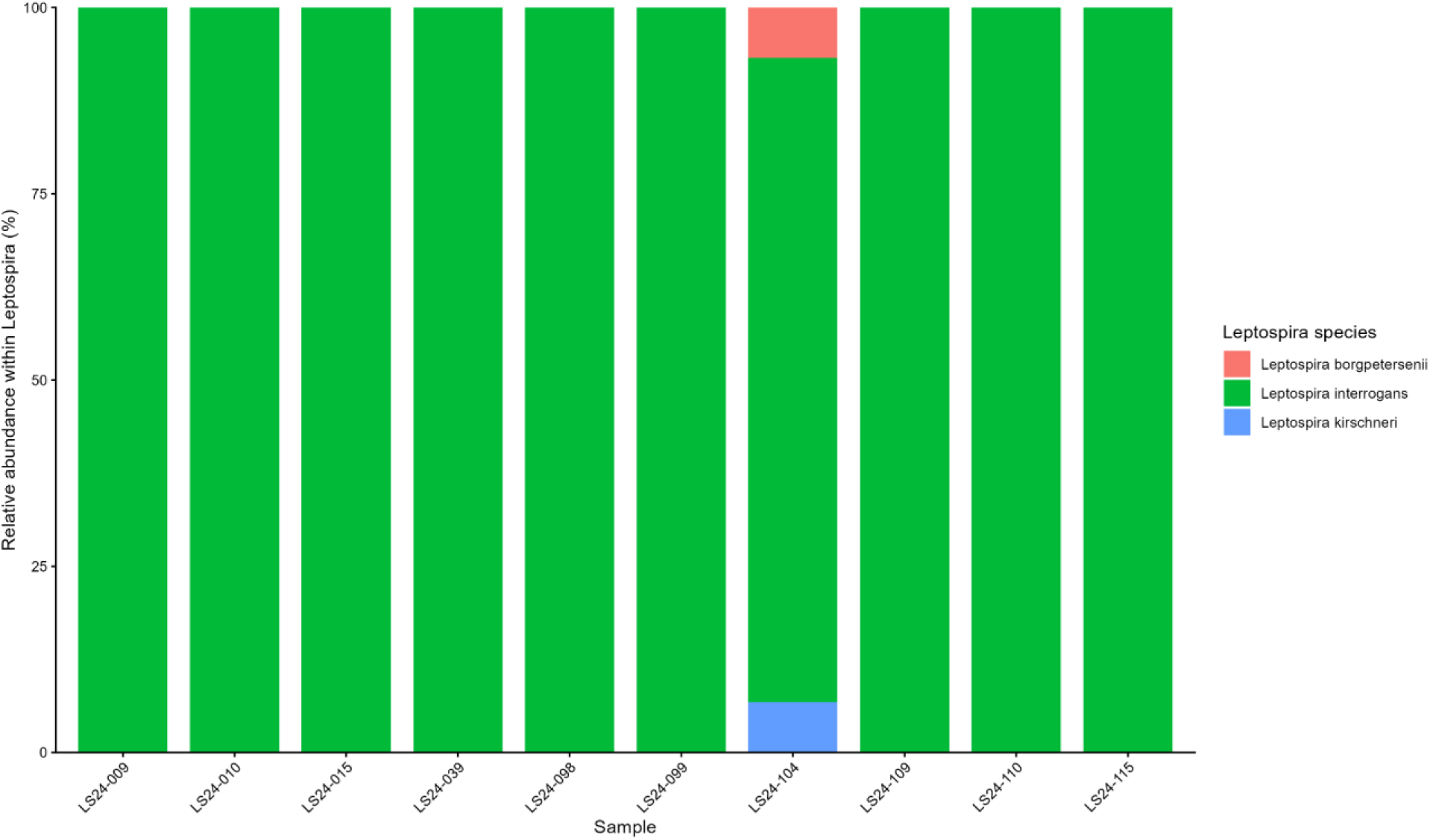
Relative abundance of *Leptospira* species among high-confidence *Leptospira* assignments

An important finding was the ability of metagenomic sequencing to provide species-level information in qPCR-positive samples that were MAT-negative. Using the filtering criteria, 60% (n=6/10) samples contained high-confidence *Leptospira* assignments, with *L. interrogans* predominating. Species-level diversity was particularly evident in LS24-104, which contained assignments to *L. interrogans, L. borgpetersenii, L. kirschneri.* This broader taxonomic resolution contrasts with the MAT results in this study, which identified reactions corresponding to *L. biflexa* serovar Patoc and *L. interrogans* serovar Autumnalis. However, these findings should not be interpreted as evidence that MAT-identified serovar was incorrect, because species-level metagenomic classification and serological serovar identification measure different biological features (Sykes et al., 2022b). MAT detects antibody responses against a panel of antigens, whereas metagenomic sequencing detects nucleic acid derived from organisms present in the specimen (Sykes et al., 2022b). Moreover, serological cross-reactivity among *Leptospira* serovars is well documented, and paradoxical reactions may result in the highest antibody titer being directed against a serovar other than that of the infecting strain (Caimi et al., 2012; Sykes et al., 2022b). The differences observed between MAT and metagenomic classification therefore underscore the complementary nature of these approaches rather than establishing a direct one-to-one correspondence between serovar and species identification (Cerqueira et al., 2010; Thaipadungpanit et al., 2011).

The clinical utility of mNGS for leptospirosis has been demonstrated in a range of challenging clinical presentations, particularly when conventional diagnostic methods are inconclusive or pathogen burden is low. mNGS has enabled diagnosis of neuroleptospirosis from cerebrospinal fluid despite negative conventional workups (Wilson et al., 2014), and has provided etiological confirmation in severe pulmonary disease and diffuse alveolar hemorrhage when cultures and serological testing were unrevealing (Chen et al., 2021; Ji et al., 2023). Its ability to detect *L. interrogans* in multiple specimen types has also facilitated diagnosis of complex co-infections, including concomitant leptospirosis and tuberculosis (Shi et al., 2022). Furthermore, cohort and case studies have shown that early mNGS diagnosis can support timely antimicrobial therapy and may be particularly valuable during the acute, antibody-negative phase of infection or after antimicrobial exposure has reduced culture yield (Jiang et al., 2022; Lu et al., 2022). Unlike these studies, which primarily demonstrate the role of mNGS in acute clinical diagnosis and treatment, our study highlights its utility for retrospective species-level detection and characterization of *Leptospira* in archived clinical specimens, especially in qPCR-positive but MAT-negative samples.

The predominance of *L. interrogans* in our metagenomic data is consistent with the molecular epidemiology of leptospirosis in the Philippines, where transmission is strongly influenced by monsoon rainfall and flooding (Matsushita et al., 2018). Several pathogenic *Leptospira* lineages have been reported in the country, including *L. interrogans* serovars Manilae and Losbanos, serogroup Grippotyphosa, and *L. borgpetersenii* serogroup Javanica (Villanueva et al., 2010). Serogroup Pyrogenes, particularly *L. interrogans* serovar Manilae, has also been associated with human and animal infections in Metro Manila (Yanagihara et al., 2007), while molecular typing of clinical isolates has identified *L. interrogans* ST12 associated with serovar Manilae as a persistent local lineage (Mendoza & Rivera, 2021). Although our shotgun metagenomic data did not provide serovar- or serogroup-level resolution, the predominance of *L. interrogans* is consistent with these previous findings and supports the potential of mNGS as a culture-independent approach for retrospective surveillance. Higher pathogen genome coverage, achieved through deeper sequencing or targeted enrichment, will nevertheless be necessary to obtain reliable strain- or serovar-level resolution from low-load clinical specimens (Lehmann et al., 2014; Grillova et al., 2023).

These findings highlight the need for strategies that improve the recovery and sequencing depth of low-abundance *Leptospira* DNA from clinical specimens. Due to the vast excess of host-derived nucleic acids in the samples, untargeted metagenomic sequencing may provide insufficient coverage of the pathogen DNA for reliable genomic characterization. Targeted enrichment or hybridization-capture offers one approach to overcome this limitation (Grillova et al., 2023). This approach may be particularly useful for qPCR-positive samples that yield few or no *Leptospira* reads by conventional metagenomic sequencing. Selective culture enrichment has likewise been shown to improve *Leptospira* recovery from samples containing competing microorganisms (Gorman et al., 2022). Integrating host depletion, targeted capture, or selective enrichment with high-throughput sequencing may therefore improve pathogen recovery and enable higher-resolution characterization while retaining the broad, culture-independent detection capability of metagenomic sequencing (Spatz & Afonso, 2024).

Several limitations should be considered when interpreting these findings. First, the study used archived serum-derived total nucleic acid extracts, which may have undergone nucleic acid degradation during storage. Second, the sequencing workflow did not incorporate host-depletion or targeted pathogen enrichment steps, potentially limiting the proportion of sequencing reads available for *Leptospira.* Finally, the relatively low pathogen abundance and limited genomic coverage precluded reliable strain- or serovar-level characterization. Future studies using prospectively collected specimens, optimized host-depletion or targeted-enrichment strategies, and greater sequencing depth may improve pathogen recovery and enable higher-resolution genomic characterization.

In summary, our findings demonstrate that metagenomic sequencing can complement established diagnostic methods for leptospirosis by providing species-level information that is not readily obtained through serology alone. High-confidence *Leptospira* assignments were recovered from a substantial proportion of qPCR-positive specimens, including MAT-negative samples, although detection remained constrained by the low abundance of pathogen DNA and substantial host-derived background. The predominance of *L. interrogans* and detection of additional pathogenic *Leptospira* species demonstrate the potential of mNGS to contribute to molecular surveillance of leptospirosis in the Philippines. At the same time, the detection of non-*Leptospira* species highlights the need for cautious interpretation of metagenomic taxonomic assignments, particularly when evaluating low-abundance organisms in clinical specimens. Overall, the recovery of species-level *Leptospira* information from archived specimens supports the feasibility of mNGS as a complementary approach to conventional diagnostics. Integrating mNGS with qPCR and serological testing, together with improved host-depletion or target-enrichment strategies, could provide a more comprehensive framework for monitoring the diversity, distribution, and epidemiology of *Leptospira* circulating in the Philippines.

## DATA SUMMARY

The R script used and the output figures and tables are available in a GitHub repository (https://github.com/lanadelrea/PH-Leptospirosis-Analyses). The FASTQ files with human host removed reads are associated with BioProject PRJNA1529875 and their corresponding SRA accession are as follows: SRR40694506, SRR40694505, SRR40694494, SRR40694493, SRR40694492, SRR40694491, SRR40694490, SRR40694489, SRR40694488, SRR40694487, SRR40694504, SRR40694503, SRR40694502, SRR40694501, SRR40694500, SRR40694499, SRR40694498, SRR40694497, SRR40694496, SRR40694495.

## ETHICAL CLEARANCE

The Research Institute for Tropical Medicine Institutional Review Board granted ethical approval for this work under IRB Assigned Number 2024-17. Patient consent was waived due to the retrospective design of the study and the use of anonymized archived DNA samples collected through the national surveillance program (PIDSR, 2018–2020). The study did not involve any patient identifiers, and all samples were labeled with laboratory-assigned codes.

## ACKNOWLEDGEMENTS

The authors would like to express sincere gratitude to all who supported this study. Special thanks to the Advanced Molecular Technologies Laboratory (AMTL) for providing the reagents critical to this research, and to the Microbiology Department for sharing the necessary samples, with particular appreciation to Gabriel Terrado, Mark Philip Bugayong, Ricaflor Banhaw and Chona Mae Daga for their contributions. We want to acknowledge Aldrin V. Imbag for providing and maintaining the high-performance computing resources that enabled the assembly and analysis of data.

## AUTHOR CONTRIBUTIONS

AMLR: Conceptualization, Methodology, Software, Validation, Formal Analysis, Investigation, Data Curation, Writing (original draft), Visualization; JIGM: Conceptualization, Methodology, Validation, Formal Analysis, Investigation, Data Curation, Writing (review and editing); LLMD: Resources, Writing (review and editing), Supervision, Project Administration, Funding Acquisition; KADC: Investigation, Resources; ANAD: Conceptualization, Methodology, Investigation, Resources, Writing (review and editing); CBAR: Investigation, Resources, Writing (review and editing); DDA: Investigation, Resources; RAB: Investigation, Data Curation, Resources; EJGM: Investigation, Resources; ADN: Investigation, Writing (review and editing); TJRD: Writing (review and editing), Supervision, Project Administration, Funding Acquisition; FGMP: Conceptualization, Methodology, Writing (review and editing), Supervision; AAPL: Conceptualization, Methodology, Writing (review and editing), Supervision, Project Administration, Funding Acquisition.

